# Smoking and socio-economic factors linked to acute exacerbations of COPD: analysis from an Asthma + Lung UK survey

**DOI:** 10.1101/2022.04.25.22274262

**Authors:** Parris J Williams, Andrew Cumella, Keir EJ Philip, Anthony A. Laverty, Nicholas S Hopkinson

**Affiliations:** National Heart and Lung Institute, Imperial College; Asthma + Lung UK; Public Health Policy Evaluation Unit, School of Public Health, Imperial College London, London, UK

**Author notes:** (Corresponding author) Professor Nicholas S. Hopkinson, National Heart and Lung Institute, Imperial College, Royal Brompton Hospital Campus, Fulham Road, London SW3 6HP, Email Address.

**Keywords:** Smoking cessation, inequality, fuel poverty, social justice, occupational exposures

## Abstract

Understanding the factors driving acute exacerbations of COPD is key to reducing their impact on human health and wellbeing. 5997 patients, mean 66 years, 64% female, completed an online survey between December 2020 and May 2021 about living with COPD developed by the charity Asthma+Lung UK. The 3731(62.2%) reporting frequent(≥2/year) exacerbations were more likely to smoke (AOR 1.70, 95%CI 1.470-1.98), have lower annual household income (≤£20,000, (AOR: 1.72, 1.36-2.17), live in a cold and damp home (AOR: 1.78, 1.50-2.11), and report previous occupational exposure to dust, fumes, and chemicals. Strategies to improve COPD outcomes must address issues of social justice.

## INTRODUCTION

Chronic obstructive pulmonary disease is highly prevalent, with at least 1.3 million people diagnosed in the UK, and rates increasing over the past decade. COPD is known to be less common in affluent populations. The gap in COPD mortality has widened dramatically in recent years, being five times higher in the lowest income decile than the highest in 2020 compared to twice as high in 2010(1) Acute exacerbations of COPD (AECOPD) in patients lead to a higher risk of hospitalisation and mortality; increased exposure to infectious pathogens, environmental pollutants and poor physical fitness are established risk factors(2). More than 50% of the cost of COPD is attributable to exacerbation management(2), so prevention is an important issue for sustainability of healthcare systems as well as for individual patients. Information about the impact of disparities in wealth and other specific housing, social and environmental factors is needed to drive efforts to address this.

## METHODS

We conducted a secondary analysis using data from an online Asthma + Lung UK annual COPD survey conducted December 2020 to May 2021, which was advertised via social media, direct email messages to the charity’s known supporter base and via its website. Full survey provided (online supplement).

### Statistical analysis

Descriptive results are presented as number (%) and mean (SD) as appropriate. Logistic regression results are presented as adjusted odds ratios (AOR) with 95% CI. Responses to the survey question “In the past 12 months, how many exacerbations or ‘flare-ups’ of your COPD symptoms have you had?” were grouped into 0-1 (infrequent) and ≥2 (frequent exacerbators), in line with current treatment guidelines. We also compared participants who did or did not report requiring hospitalisation for an AECOPD in the preceding year. All regression analyses included age, gender, and smoking status as independent covariates. Ex-smokers, no previous occupational exposure to airborne pollutants, warm and dry housing, and Household income ≥£40,000 were used as reference categories for the multiple regression analyses.

## RESULTS

The initial sample included 8,232 responses. After cleaning for outliers, removing duplicate and incomplete responses, 5997 responses remained and were analysed. The sample population was majority female (64.4%), white ethnicity (99%) and ex-smokers (80.7%). Mean age was 66.2(8.9) years.

The 3731 frequent exacerbators were more likely to be current smokers (AOR: 1.70, 95% CI 1.47-1.98), have low annual household (HH) incomes, (≤£20,000 (AOR: 1.72, 95%CI, 1.36-2.17), live in cold and damp housing (AOR: 1.78, 95% CI 1.50-2.11), report previous occupational exposure to airborne pollutants (AOR:1.12, 95% CI 1.00-1.25) and be male (AOR: 1.32, 95%CI, 1.17-1.49) (Table 2, Figure 1).

**Table 1.**
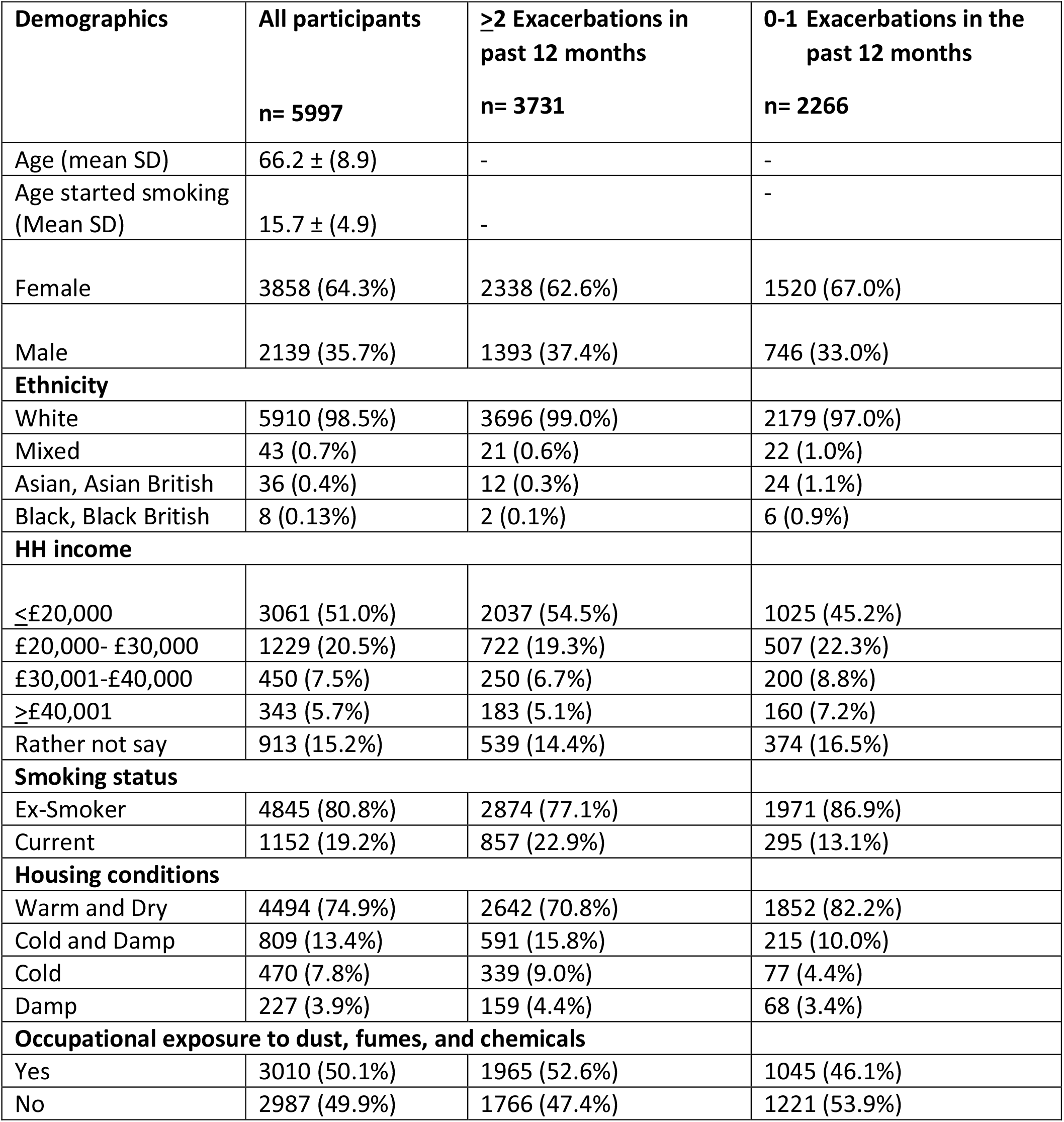
Sociodemographic characteristics of the survey respondents sample including percentage of the sample demographics in each exacerbation group.

**Table 2.** Factors associated with increased number of exacerbations among survey population.

|  | **Model 1: age, sex tobacco & income** | **Model 2: age, sex, tobacco & housing** | **Model 3: All Variables** |
| --- | --- | --- | --- |
| **Variable** | **OR and CI** | **OR and CI** | **OR and CI** |
| Gender Female | Ref | Ref | Ref |
| Male | **1.38 (1.23-1.54)** | **1.32 (1.18-1.48)** | **1.32(1.17-1.49)** |
| Age | **0.97 (0.96-0.97)** | **0.97 (0.96-0.98)** | **0.97 (0.96-0.98)** |
| Smoking status (current) | **1.68 (1.45-1.95)** | **1.77 (1.53-2.06)** | **1.70 (1.47-1.98)** |
| Ex-Smokers | Ref | Ref | Ref |
| Occupational Exposure to dust, fumes, and Chemicals | - | - | **1.12 (1.00-1.25)** |
| No occupational exposure to dust, fumes, and chemicals | - | - | Ref |
| Age started Smoking | - | - | 1.001(0.990-1.012) |
| Housing: Warm and Dry | - | Ref | Ref |
| Housing: Cold and Damp | - | **1.83 (1.54-2.17)** | **1.78(1.50-2.11)** |
| Housing: Cold | - | **1.69 (1.37-2.09)** | **1.61 (1.30-2.00)** |
| Housing Damp | - | **1.52 (1.13-2.05)** | **1.49 (1.01- 2.00)** |
| HH income: ≤£20,000 | **1.82 (1.45-2.30)** | - | **1.72 (1.36-2.17)** |
| HH income: £20,001-£30,000 | **1.32 (1.03- 1.68)** | - | **1.27 (0.99- 1.63)** |
| HH income: £30,001-£40,000 | 1.11 (0.83-1.48) | - | 1.08 (0.81-1.44) |
| HH income: ≥£40,000 | Ref | - | Ref |
| HH Income: Rather not say | **1.46 (1.13-1.88)** | - | **1.40 (1.08-1.82)** |

*Bold type indicates significance at ≤0.05 level*

**Figure 1.**
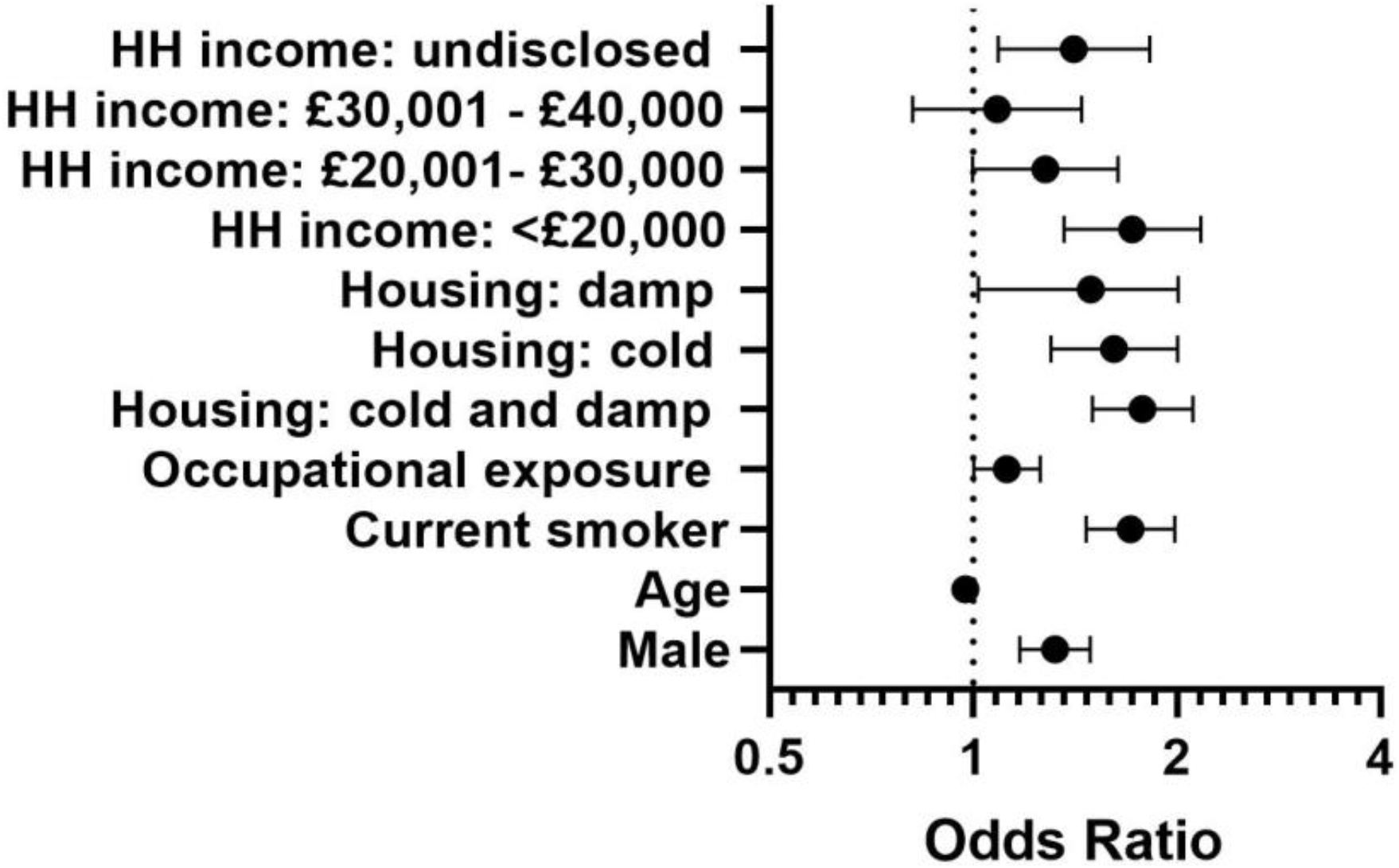
Smoking, socio-economic and housing factors affecting frequency of AECOPD among survey population (Adjusted odds ratios, 95% CI).

Among the 532(8.9%) of survey respondents who reported a hospital attendance to manage their most recent AECOPD, 117(22%) were current smokers. Smokers were more likely to report hospitalisation compared to ex-smokers (AOR: 1.25, 95% CI 0.99-1.59) Both low income and cold and damp housing covariates were associated with numerically higher in those reporting hospitalisation, but these differences were not statistically significant (Online Supplement).

## DISCUSSION

This large online survey of the experience of people living with COPD, provides important contemporary data on the role that socioeconomic factors may play in AECOPD. We found that low income, poor housing quality, past occupational exposure to airborne pollutants and current smoking were all significantly associated with higher AECOPD frequency. Current smoking was also associated with an increased risk of AECOPD requiring hospitalisation.

A key objective in COPD care is to reduce the frequency of AECOPD, both to improve patients’ quality of life and to limit as far as possible the avoidable use of finite healthcare resources. However, there is a huge unmet care need among COPD patients within the UK, with a large proportion of patients missing out on important COPD care such as self-management plans, vaccinations, pulmonary rehabilitation and smoking cessation(3), all of which are proven interventions for reducing AECOPD. The COVID-19 pandemic has, worsened this unmet need further, as people with respiratory disease have had trouble accessing healthcare(4, 5). Asthma+Lung UK survey data show that over 75% COPD patients report not receiving basic care during 2020/2021(6). Switching to a digital by default model, further risks excluding deprived and older patients.

Despite the known link between housing quality and health (particularly excess winter deaths), and the 2015 National Institute for Health Care and Excellence(NICE) guidance which recommends that healthcare providers assess housing quality and make referrals where necessary(8), the effect of housing quality on COPD health has been little studied. Current guidance for housing temperature in the winter is a minimum of 18°C(8) but, this may not be sufficient for people with COPD. A 2008 paper reported that greater time spent with an indoor temperature ≥21°C was associated with better self-reported health status in people with COPD(9). Of note, data from the Office for Health Improvement and Disparities(OHID) show that in 2018 2.4 million people in England were living in fuel poverty(1). Most survey participants fell below the median household income in UK, which is £31,004, and the link between the subjective experience of cold and/or damp linked to AECOPD in part reflects that, though it was also associated independent of income.

Cutting social support through austerity policies, has affected the most disadvantaged COPD populations, contributing to the structural violence that people with the condition experience(7). Our findings corroborate this, with the poorer survey respondents almost twice as likely to be in the frequent exacerbator groups and are especially relevant as fuel poverty increases.

The survey highlights an important link between smoking, AECOPD frequency and hospitalisations, adding further urgency to the need for strategies to deliver the UK government’s smokefree2030 ambition(10).

The survey design has some limitations, requiring digital literacy which may limit generalisability, but it adds to the growing evidence that socioeconomic status, in particular poor housing conditions, are linked with increased frequency of AECOPD.

Addressing social deprivation within this population is essential to reduce inequalities and reduce the poor value intrinsic in needing to provide healthcare for acute conditions rather than preventing them.

## Supporting information

Full survey provided (online supplement).

## Data Availability

Anonymised research data will be shared with third parties via request to Asthma + Lung UK.

## DATA SHARING

Anonymised research data will be shared with third parties via request to Asthma + Lung UK.

## ETHICS STATEMENT

Ethical approval was granted by the Imperial College Research Governance and Integrity Team (ICREC Ref: 20IC6625).

## AUTHOR STATEMENT

NSH, PJW, AC, AL designed the study. PJW and AL analysed the data and PJW produced the first draft to which all authors contributed. All authors have reviewed and approved the final version. NSH is the guarantor.

## TRANSPARENCY DECLARATION

Nicholas Hopkinson, the manuscript’s guarantor affirms that the manuscript is an honest, accurate, and transparent account of the study being reported; that no important aspects of the study have been omitted; and that any discrepancies from the study as planned (and, if relevant, registered) have been explained

