## Supplementary material for "Smoking and socio-economic factors linked to acute exacerbations of COPD: analysis from an Asthma + Lung UK survey": Full survey provided (online supplement).

Parris J Williams[1], Andrew Cumella[2], Keir EJ Philip[1], Anthony A. Laverty[3], Nicholas S Hopkinson[1].

[1]National Heart and Lung Institute, Imperial College

[2]Asthma + Lung UK

[3]Public Health Policy Evaluation Unit, School of Public Health, Imperial College London, London, UK

Page 2 Additional methods

Page 4 Factors associated with hospitalisation for AECOPD

Page 5 Survey questions

### Additional Methods

**Model 1: Household income**

Table 1. Smoking and annual household income factors associated with increased AECOPD

| Variable | Odds Ratio | 95%CI | P Value |
|---|---|---|---|
| **Gender (Male)** | 1.38 | 1.23-1.54 | 0.00 |
| **Smoking** | 1.68 | 1.45-1.95 | 0.00 |
| **Age** | 0.97 | 0.96-0.97 | 0.00 |
| **< £20,000** | 1.82 | 1.45-2.30 | 0.00 |
| **£20,001- £30,000** | 1.32 | 1.03- 1.68 | 0.004 |
| **£30,001- £40,000** | 1.11 | 0.83-1.48 | 0.14 |
| **More than £40,000** | Ref | Ref | Ref |
| **Rather not say** | 1.46 | 1.13-1.88 | 0.00 |

In the above regression model, we categorised annual household income into 5 groups; group 1: <£20,000, group 2: £20,000- £30,000, group 3: £30,001- £40,000, group 4: rather not say and group 5: more than £40,000. The reference category we used in this model was group 5 (>£40,000). Survey respondents were more likely to report higher AECOP frequency if they were male (OR 1.138, 95%CI 1.23-1.54, p= 0.00), current smokers (OR: 1.68, 95%CI 1.45-1.95), and had lower annual HH incomes <£20,000 (OR: 1.82, 95%CI 1.45-2.30, p= 0.00), £20,001- £30,000 (OR: 1.32, 95%CI 1.03- 1.68, p= 0.004) or didn't disclose their income (OR: 1.46, 95%CI 1.13-1.88, p=0.00).

**Model 2: Housing Quality**

Table 2. Smoking and housing quality factors associated with increased AECOPD

| Variable | Odds ratio | 95%CI | P-value |
|---|---|---|---|
| **Gender (male)** | 1.324 | 1.183-1.483 | 0.00 |
| **Age** | 0.975 | 0.969-0.981 | 0.00 |
| **Smoking** | 1.779 | 1.531-2.066 | 0.00 |
| **Good Housing** | Ref | Ref | Ref |
| **Cold and Damp housing** | 1.833 | 1.547-2.171 | 0.00 |
| **Cold housing** | 1.695 | 1.370-2.098 | 0.00 |
| **Damp Housing** | 1.528 | 1.138-2.053 | 0.005 |

In this model survey respondents were more likely to report >2 exacerbations over the past year if they were male (OR: 1.324, 95% CI 1.183-1.483 p= 0.00) and were a current smoker (OR: 1.778, 95% CI 1.531-2.066 p= 0.00). To observe if housing conditions influenced exacerbations we used 'Good Housing' as the reference category in this model. Survey responders were more likely to report more than 2 exacerbations in the past year if they lived in cold and damp housing (OR: 1.833, 95%CI 1.547-2.171 p-0.00), cold housing (OR: 1.695, 95%CI 1.370-2.098 p = 0.00) and damp housing (1.528, 95% CI 1.138-2.053 p =0.005) compared to living in good housing conditions.

**Frequency of AECOPD grouped.**

For the original analysis we grouped AECOPD frequencies into two categories 1= 0-1 AECOPD and 2= ≥2 AECOPD. The graph below gives a visual representation of the frequencies of AECOPD collapsing ≥6 into one group.

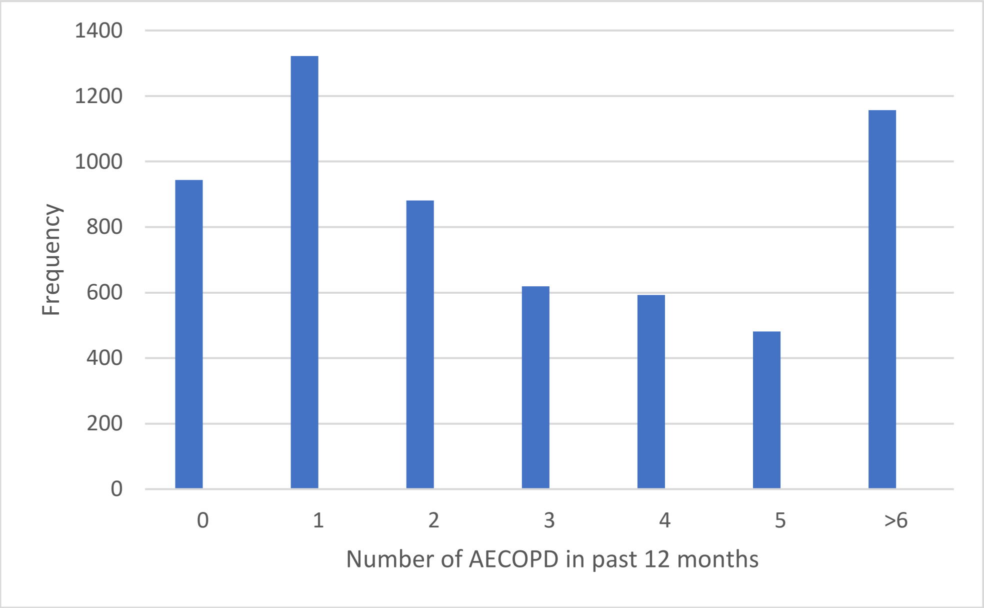


*Figure 1. Frequencies of AECOPD in the past 12 months in sample population.*

### Smoking and socio-economic factors associated with hospitalisations of AECOPD

We ran another logistic regression model to determine if smoking and socio-economic factors my contribute to hospitalisations of AECOPD within the survey population. We took the survey question "how did you manage your most recent flare up?" and coded it into two binary numeric categories 0= No hospitalisation 1= hospitalisation. Patients reporting 0 exacerbations were excluded from the analysis.

Table 1. Smoking and socio-economic factors associated with attendance to hospital to manage recent AECOPD.

| **Variable** | **Odds Ratio** | **95%CI** | **P vale** |
|---|---|---|---|
| **Gender (Male)** | 0.94 | 0.77- 1.14 | 0.53 |
| **Age** | 0.99 | 0.99-1.00 | 0.70 |
| **Smoking (current)** | 1.25 | 0.99-1.59 | 0.05 |
| **Age started** | 1.00 | 0.99-1.02 | 0.40 |
| **Housing: Warm and Dry** | Ref | Ref | Ref |
| **Housing: cold and damp** | 1.08 | 0.84-1.40 | 0.50 |
| **Housing: Cold** | 0.95 | 0.68-1.33 | 0.78 |
| **Housing: Damp** | 0.74 | 0.455-1.22 | 0.24 |
| **Occupational Exposure of dust, fumes or chemicals** | 1.04 | 0.87-1.25 | 0.63 |
| **<£20,000** | 1.13 | 0.76-1.73 | 0.53 |
| **£20,001- £30,001** | 0.93 | 0.63-1.44 | 0.75 |
| **£30,001- £40,000** | 0.55 | 0.31-1.004 | 0.05 |
| **>£40,000** | Ref | Ref | Ref |
| **Rather not say** | 1.12 | 0.70-1.72 | 0.66 |

Survey respondents were more likely to report a hospital admission to manage their AECOPD if they were current smokers (AOR: 1.25, 95%CI 0.99-1.59, p=0.05), HH incomes >£20,000, occupational exposure to airborne pollutants, living in a cold and damp house and age started smoking we also covariates moderately associated with hospitalisations, however not statically significant. Interestingly survey respondents reporting their annual HH income as £30,001-£40,001 were less likely to report attending hospital compared to those reporting their annual HH income as >£40,000 (AOR 0.55, 95%CI 0.31-1.00, p= 0.05).

Survey questions.

Further results are available in the Failing on the Fundamentals report from Asthma + Lung UK can be found Here the questions asked in the survey to gain the data usDataed in this report.

1. What is your age?
   [respondent typed in numerical value]

2. What nation do you live in?
   • England
   • Northern Ireland
   • Scotland
   • Wales

3. [for respondents in England] Which region do you live in?
   • East Midlands
   • East of England
   • London
   • North East
   • North West
   • South East
   • South West
   • West Midlands
   • Yorkshire and Humber

4. When were you diagnosed with COPD?
   • In the last 6 months
   • 6 months – a year ago
   • 1 year – 2 years ago
   • 2 years – 5 years ago
   • 5 years – 10 years ago
   • Over 10 years ago
   • I am waiting for my diagnosis

5. How long did you experience COPD symptoms before talking about it with your GP or another healthcare professional?
*Common symptoms include increasing breathlessness, a persistent chesty cough with phlegm that does not go away, frequent chest infections and persistent wheezing.*
• One month or less

• 1-2 months

• 3-6 months

• 6 months – a year

• A year – two years

• More than two years

• I don’t remember

6. How long did you have to wait between talking about your COPD symptoms with your GP or healthcare professional, to receiving a formal diagnosis?
• One month or less

• 1-2 months

• 3-6 months

• 6 months – a year

• A year – two years

• More than two years

• I don’t remember

7. What were the main barriers to getting a diagnosis?
• Difficulty in getting appointments

• Diagnosis tests (such as spirometry) not being available

• Not knowing what the signs of potential COPD were

• Not wanting to know if I had COPD

• I was misdiagnosed at first, and it took a while to get correctly diagnoses with COPD

• I was sent away by my GP when I first mentioned it

• My GP thought I had a chest infection or cough at first

• I do not recall there being any barriers

• Concern that I might have lung cancer

• Other

8. Thinking about when you were diagnosed, did you have any of the following performed to confirm your diagnosis?
   • Spirometry testing

   • A peak flow reading

   • A chest x-ray

   • A CT scan

   • Blood tests

   • I was asked about smoking

   • We had a discussion about smoking

   • We had a discussion about my symptoms

   • We had a discussion about my work history

   • I don't remember

   • None of the above

9. After diagnosis, were you given any written materials to support your management of your COPD? This could include leaflets or links to the BLF website, for instance.
   • Yes

   • No

   • Don't remember

10. After your diagnosis, did you feel you had enough knowledge and support to manage your COPD?
   • Yes

   • No

   • Don't remember

11. Are you currently working?
   • Yes

   • No

12. Have you worked in a job where you were exposed to dust, fumes or chemicals? This may include working in a factory, using cleaning products or working in farming.
   • Yes

- No

13. [if yes to above] In the job(s) where you were exposed to dust, fumes or chemicals, do you think that you had all the protective equipment that you needed to stay safe?
    - Yes
    - No

14. [if yes to working with dust etc] Do you think your job made your condition worse?
    - Yes
    - No

15. When do you get out of breath?
    - I'm not troubled by being out of breath, except on strenuous exercise
    - I'm short of breath when hurrying on level ground or walking up a slight hill
    - I walk slower than most people on the level, stop after a mile or so, or stop after 15 minutes of walking at my own pace
    - I stop for breath after walking about 100 yards or after a few minutes on level ground
    - I'm too breathless to leave the house, or breathless when dressing and undressing

16. Have you had a planned review or planned check-up (sometimes called an annual review) of your COPD with your doctor or nurse in the last year?
    - Yes - it was done face to face
    - Yes - it was done over the phone / via videocall
    - Yes - it was done via text
    - No
    - Not sure

17. Do you smoke?
    - Yes
    - I used to, but have given up
    - I have never smoked

18. How old were you when you had your first cigarette?

    [respondent typed in numerical value]

19. [for those who used to smoke] Did you give up smoking after being diagnosed with COPD?
    - Yes

- No

20. [for those who smoke, or have smoked in past 12 months] In the past 12 months, have you been offered treatment and support to stop smoking?

    - Yes
    - No

21. [current smokers or used to] Since being diagnosed with COPD, have you tried to stop smoking?

    - Yes
    - No
    - I wasn't smoking at the time I was diagnosed

22. What inspired you to quit smoking?

    - Being diagnosed with COPD
    - Other health reasons
    - Stop Smoking campaigns (such as Stoptober)
    - My family and/or friends
    - Protecting others
    - Saving money
    - Other

23. In the past 12 months, have you had a flu jab?

    - Yes
    - No
    - Don't know

24. Since diagnosis with COPD, have you had a pneumonia vaccine jab? This is also called the pneumococcal vaccine, or PPV.
    - Yes
    - No
    - Don't know

25. Have you had pulmonary rehabilitation as part of your care?
    *Pulmonary rehabilitation (PR) is a programme of exercise and education designed for people living with COPD and other respiratory conditions*

    - Yes
    - No

26. [if no to above] Have you ever been offered the chance to do pulmonary rehabilitation?

    - Yes
    - No

27. [if yes to PR] Has doing pulmonary rehabilitation improved your COPD symptoms?
    - Yes
    - No
    - I don't know

28. [if yes to doing PR] Would you recommend pulmonary rehabilitation to others with COPD?

    - Yes
    - No
    - I don't know

29. Do you currently have a COPD self-management plan?
    - Yes
    - No
    - Don't know

30. [if yes to above] Did you have a chance to have a say about what was in the self-management plan? This could have been done via a conversation with your healthcare professional
    - Yes
    - No
    - I can't remember

31. In the past 12 months, have you discussed any other long term medical conditions that you have in relation to your COPD management with your doctor or nurse?
    - Yes
    - No
    - Don't know
    - I don't have any other long term conditions

32. In the past 12 months, how many exacerbations or 'flare-up' of your COPD symptoms have you had?
    *By this, we mean you suffered from some of these warning signs:*
    *• Your breathlessness gets worse, and this goes on for some time without getting better*
    *• You cough more*
    *• You produce more sputum*
    • There's a change in the colour and consistency of your sputum

    [respondent typed in numerical value]

    - how did you manage your most recent flare up?
        I. I managed it myself without informing a healthcare professional
        II. I managed it myself and informed a healthcare professional
        III. I was treated at home by a GP or paramedic
        IV. I was treated over the phone e.g. NHS 111
        V. I went to hospital/ A&E by myself

VI. I went to hospital/ A&E in an ambulance
VII. I went to my GP
VIII. I went to my pharmacist

33. Do you know what to do if your COPD symptoms get worse (you have a flare-up)?

    - Yes
    - No

34. How would you rate the general public's understanding of what COPD is?
    - Very good
    - Good
    - Average
    - Poor
    - Very poor

35. How would you rate the understanding of living with COPD from the following groups:
    - Your friends and family
        i. Very good
        ii. Good
        iii. Average
        iv. Poor
        v. Very poor
    - Healthcare professionals
        i. Very good
        ii. Good
        iii. Average
        iv. Poor
        v. Very poor
    - The general public
        i. Very good
        ii. Good
        iii. Average
        iv. Poor
        v. Very poor
    - The media
        i. Very good
        ii. Good

iii. Average

iv. Poor

v. Very poor

36. What activities have you had to do less of because of your COPD? [multiple choice]

- Work
- Seeing friends
- Seeing family
- Volunteering
- Doing childcare
- Doing other sorts of care for other family or friends
- Going on holiday
- Other [free text]

37. What activities have you had to stop doing because of your COPD? [multiple choice]
    - Work
    - Seeing friends
    - Seeing family
    - Volunteering
    - Doing childcare
    - Doing other sorts of care for other family or friends
    - Going on holiday
    - Other [free text]

38. Is your COPD affected because where you live is:
    - Cold?
    - Damp?
    - Both cold and damp?

39. Have you ever felt that there is a stigma attached to living with COPD?
    - Yes
    - No
    - Don't know

40. Do you feel you have faced any stigma or discrimination due to having COPD?
    - Yes
    - No

41. [if yes to above] Can you describe what took place?

[free text]

42. How has being diagnosed with and living with COPD impacted upon your mental health?

- It has had no impact
- It has made it much better

- It has made it a bit better
- It has made it a bit worse
- It has made it much worse

43. Which of the following applies to care for your mental health since you were diagnosed with COPD? [multiple choice]
    - I have spoken to my family and/or friends about it
    - I have spoken to my GP about it
    - I have been diagnosed with a new mental health condition
    - I have received a new prescription for my mental health
    - I have been referred to a specialist for mental health treatment
    - I have not needed mental health care
    - Other
